# Genomic insights into the impact of typhoid conjugate vaccine introduction and the COVID-19 pandemic on *Salmonella* Typhi and Paratyphi A in Nepal

**DOI:** 10.64898/2026.07.21.26358582

**Authors:** Kesia Esther Da Silva, Shiva Ram Naga, Nishan Katuwal, Sabin Bikram Shahi, Denise O Garrett, Isaac I. Bogoch, Kate Doyle, Stephen P Luby, Rajeev Shrestha, Dipesh Tamrakar, Jason R. Andrews

**Author notes:** **Correspondence:** Jason R. Andrews, Division of Infectious Diseases and Geographic Medicine Stanford University School of Medicine, Biomedical Innovations Building, Room 3458 Stanford, CA 94025, USA.

## Abstract

**Background:** Enteric fever, caused by *Salmonella enterica* serovars Typhi and Paratyphi A, remains a major concern in low-and middle-income countries, with treatment increasingly complicated by antimicrobial resistance. Typhoid conjugate vaccines (TCVs) offer a promising intervention, but their impact on circulating lineages and population structure remains poorly understood. Nepal introduced a nationwide TCV programme in April 2022, shortly after the COVID-19 pandemic disrupted healthcare delivery and surveillance.

**Methods:** We sequenced 350 *S*. Typhi and 114 *S*. Paratyphi A isolates collected in Nepal between 2018 and 2024 and analyzed them alongside1,796 previously published Nepal genomes (2005-2018). We characterized genotype distribution, antimicrobial resistance determinants, phylogenetic relationships and lineage-specific phylodynamics across three epidemiological periods: pre-pandemic (before 2020), pandemic/pre-vaccine (2020–2022), and post-vaccine introduction (2022–2024).

**Results:** Sixteen *S*. Typhi genotypes were identified, dominated by 4.3.1.2 (28%), 3.3.2 (20.3%) and 3.3.1 (16.9%); *S*. Paratyphi A was dominated by genotypes 2.4.3 (57.0%) and 2.4.2 (25.4%). Multidrug resistance was rare, but fluoroquinolone non-susceptibility was widespread (70.6% of *S*. Typhi; 99.1% of *S*. Paratyphi A), with high-level resistance confined to a single *S*. Typhi 4.3.1.2.1 clade dating to 2008. Phylodynamic reconstruction revealed asymmetric lineage trajectories: *S*. Typhi H58 contracted sharply during the pandemic, lineage 3.3 contracted specifically post-TCV, and *S*. Paratyphi A (not vaccine-targeted) showed only a modest pandemic-era decline. Post-TCV, the age distribution of cases shifted older and the adult genotype mix shifted toward lineage 3.3. We found no evidence of vaccine-driven escape at the Vi capsule or O-antigen in Nepal or across a cross-country cohort.

**Conclusions:** Both *S*. Typhi and *S*. Paratyphi A populations in Nepal underwent recent declines in effective population size, coinciding with the COVID-19 pandemic and TCV introduction. Persistent fluoroquinolone resistance, particularly among *S*. Paratyphi A, underscores ongoing AMR challenges and highlights the need for expanded vaccine strategies targeting both pathogens.

**Author Summary:** Enteric fever, caused by the bacteria *Salmonella* Typhi and Salmonella Paratyphi A, remains a major cause of illness across South Asia, and its treatment is increasingly threatened by antibiotic resistance. Typhoid conjugate vaccines (TCVs) can prevent typhoid, but they do not protect against Paratyphi A, and their effect on the bacterial populations circulating in a community is not well understood. Nepal introduced TCV nationwide in 2022, shortly after the COVID-19 pandemic disrupted daily life and healthcare. We sequenced the whole genomes of *S*. Typhi and Paratyphi A collected in Nepal between 2018 and 2024 and combined them with previously published genomes to reconstruct how these populations changed over time. Different lineages declined at different times: one major typhoid lineage shrank during the pandemic, whereas another declined specifically after vaccine introduction. *S*. Paratyphi A, which the vaccine does not target, changed little and served as a natural comparator. Resistance to fluoroquinolone antibiotics remained widespread. Our findings illustrate how vaccination and large-scale social disruption can reshape the populations that cause enteric fever, and they support expanding vaccines to also cover *S*. Paratyphi A.

## Introduction

Enteric fever, caused by *Salmonella enterica* serovars Typhi and Paratyphi, is a systemic bacterial infection transmitted through the fecal-oral route and remains a major public health concern in low-and middle-income countries [1]. In 2019, an estimated 9.2 million cases of typhoid fever and approximately 110,000 associated deaths occurred worldwide, with the highest burden occurring in South Asia [2]. The persistence of enteric fever is closely linked to inadequate sanitation, unsafe water, and rapid urbanization in endemic regions [3]. In recent decades, the growing emergence and global dissemination of antimicrobial-resistant (AMR) *Salmonella* strains has further complicated disease management by reducing treatment efficacy, increasing healthcare costs, and contributing to higher morbidity and mortality [4,5].

Typhoid conjugate vaccines (TCVs) represent a major advance in the prevention of typhoid fever, offering durable immunity and suitability for use in infants and young children [6,7]. Early field evaluations have shown substantial reductions in disease incidence following vaccine introduction [8–10]. However, the broader impact of vaccination on the population structure, transmission dynamics, and antimicrobial resistance profiles of circulating *Salmonella* lineages remains poorly understood. In particular, TCVs target *S.* Typhi but do not protect against *S.* Paratyphi A, raising concerns about potential shifts in serovar distribution or lineage replacement following vaccine implementation [11].

Nepal is a high-burden setting for enteric fever, with both *S*. Typhi and *S*. Paratyphi A circulating endemically [12,13]. Previous genomic studies have shown the dominance of the globally disseminated S. Typhi genotype 4.3.1 (H58 lineage), which is frequently associated with reduced fluoroquinolone susceptibility and other antimicrobial resistance determinants [14,15]. In 2022, Nepal introduced a nationwide TCV campaign targeting children aged 15 months to 15 years, representing the first large-scale intervention against typhoid in the country [2,10]. This public health intervention occurred shortly after the COVID-19 pandemic, which had substantially disrupted healthcare access, disease surveillance systems, and population mobility, potentially altering enteric fever transmission dynamics [10].

Understanding how these concurrent events have shaped the genomic epidemiology of typhoidal *Salmonella* is critical for informing long-term disease control strategies. The close temporal proximity of COVID-19-related disruptions and the rapid nationwide introduction of TCVs created a unique epidemiological setting in which lineage-specific genomic trajectories may provide insights into the relative contributions of pandemic-associated and vaccine-associated effects on enteric fever transmission. To investigate these dynamics, we combined prospective genomic surveillance with historical genomic datasets to characterize changes in the population structure, antimicrobial resistance profiles, and transmission dynamics of *S.* Typhi and *S.* Paratyphi A in Nepal. Using whole-genome sequencing and phylodynamic analyses, we evaluated trends in circulating genotypes and effective population size before the COVID-19 pandemic, during the pandemic period, and following TCV introduction.

## Methods

### Study design

We conducted hospital based surveillance between 2018 to 2024 using two modalities; prospective surveillance and retrospective surveillance. In prospective surveillance, we prospectively enrolled patients presenting to outpatient or emergency departments with fever of ≥3 consecutive days’ duration within the preceding 7 days, and residing within the predefined catchment area and performed the blood culture. In addition, patients admitted to inpatient wards with suspected or confirmed enteric fever were also enrolled. Prospective surveillance was implemented at three tertiary-care hospitals: Dhulikhel Hospital, Kathmandu University Hospital (DHKUH) (2018-2024) in Kavrepalanchok district, Kathmandu Medical College Teaching Hospital (2018-2020) in Kathmandu district, and Kathmandu Model Hospital (2022-2024) in Kathmandu district. In retrospective surveillance, we enrolled all the blood culture confirmed enteric fever cases from hospital laboratories. This was conducted at hospital laboratory of prospective site as well as additional hospitals, including Bir Hospital (2018-2024), Nepal Medical College (2018-2022), Helping Hands Community Hospital (2018-2022), Kathmandu Model Hospital (2018-2022), Alka Hospital (2018-2020), Kanti Children’s Hospital (2019, 2023-2024), Nepal Police Hospital (2018-2019), Siddhi Memorial Women and Children Hospital (2022-2024), and Civil Service Hospital (2022-2024).

### Bacterial isolates

Whole-blood specimens were collected aseptically and incubated using the BACTEC automated blood culture system (Becton Dickinson, Franklin Lakes, NJ, USA). Bacterial identification was performed using standard biochemical methods, and serological confirmation was carried out using O and H antisera (MAST ASSURE, Mast Group, Liverpool, UK), where available. Isolates obtained from the hospital laboratories and the retrospective laboratory network sites were reconfirmed at DHKUH using the same biochemical and serological procedures to ensure consistency.

### Whole genome sequencing

All available *S*. Typhi and *S*. Paratyphi A isolates collected between 1 February 2018 and 27 February 2024 were included for whole-genome sequencing. These newly sequenced isolates obtained through both prospective and retrospective enrolment were analysed together with previously published Nepal genomes that provide historical and phylogenetic context. Genomic DNA was extracted from bacterial cultures using the Qiagen QIAamp DNA Mini Kit (Qiagen, Hilden, Germany) following the manufacturer’s instructions. Whole-genome sequencing libraries were prepared and sequenced on the Illumina NovaSeq X Plus platform (Illumina, San Diego, CA, USA) generating paired-end reads (2 × 150 bp). Sequence quality was assessed using FastQC v0.12.1 [16], and quality metrics were summarized using MultiQC v1.31 [17]. Species identification was confirmed using Kraken2 [18]. De novo genome assemblies were generated using SPAdes v3.15.2 [19].

### Mapping and single-nucleotide polymorphism (SNP) analysis

Paired-end Illumina reads were mapped to the *S*. Typhi CT18 (accession no. AL513382) reference chromosome sequence using RedDog mapping pipeline v1beta.11. Chromosomal SNPs with confident homozygous calls (Phred score >20) present in more than 95% of genomes were retained to define a soft core genome. SNPs were concatenated to generate a multiple sequence alignment using the RedDog script parseSNPtable.py. SNPs located within prophage regions and repetitive sequences of the CT18 reference genome were excluded. Recombination events were detected using Gubbins v2.4.1 [20] and removed from the alignment. *S*. Typhi genotypes were called directly from Illumina reads using Mykrobe v0.13.0 [21] with the Typhi typing panel (v20221207) and summarized using the GenoTyphi pipeline (https://github.com/typhoidgenomics/genotyphi). For *S*. Paratyphi A, a similar analytical pipeline was applied using the AKU_12601 reference genome (accession number FM200053). Genotypes were assigned using the Paratype genotyping framework [22]. To provide global context, additional *S.* Typhi and *S*. Paratyphi A genomes from Nepal [14,15,23–27] were downloaded from the European Nucleotide Archive and subjected to the same SNP–calling and recombination filtering pipeline.

### Antimicrobial resistance analysis

Antimicrobial resistance determinants were identified using Short Read Sequence Typing 2 (SRST2) (v0.2.0) [28] to screen sequencing reads against the ARGannot database [29] for resistance genes and the PlasmidFinder database [30] for plasmid replicons. Isolates were classified as multidrug resistant (MDR) if resistance genes associated with β-lactams, trimethoprim, sulfonamides, and chloramphenicol were detected. Point mutations within the quinolone resistance-determining regions (QRDRs) of the *gyrA*, *gyrB*, *parC*, and *parE* genes were identified using Mykrobe v0.13.0, along with plasmid-mediated quinolone resistance genes such as *qnrS*. Mutations in the *acrB* gene associated with azithromycin resistance were also detected using Mykrobe v0.13.0. Antimicrobial susceptibility was inferred genotypically from these resistance determinants and QRDR mutations.

### Phylogenetic analysis

Maximum likelihood (ML) phylogenetic trees were inferred from the chromosomal SNP alignments using RAxML v8.2.10, command raxmlHPC-PTHREADS [31]. A generalized time-reversible model and a gamma distribution were used to model site-specific rate variation (GTR+ Γ substitution model; GTRGAMMA in RAxML) with 100 bootstrap pseudo-replicates used to assess branch support for the ML phylogeny. We selected the single tree with the highest likelihood score as the best tree. The resulting phylogenies were visualized and annotated using the iTOL v7 online version [32].

### Temporal phylogenetic analysis

To investigate the evolutionary history of the major typhoidal *Salmonella* lineages circulating in Nepal, Bayesian temporal phylogenetic analyses were conducted for the dominant *S*. Typhi and *S*. Paratyphi A lineages. For the H58 lineage (genotype 4.3.1), including its sublineages 4.3.1.1 and 4.3.1.2, a total of 359 genomes from our dataset and previously published studies were selected by stratified random sampling across sampling years and included in the analysis, spanning a broad temporal range in order to capture the long-term evolutionary history of this lineage. For *S*. Typhi lineage 3.3 (including genotypes 3.3.1 and 3.3.2), 376 isolates were analyzed, representing all lineage 3.3 genomes available in our dataset. For *S*. Paratyphi A, all isolates from the full Nepal dataset (n = 538; 114 newly sequenced and 424 previously published) were included in the temporal phylogenetic analysis.

Time-calibrated phylogenies were inferred using a Bayesian Markov chain Monte Carlo (MCMC) framework implemented in BEAST2 v2.6.2 [33]. Sampling times were incorporated as tip dates corresponding to the year of isolation. Model testing indicated that a generalized time-reversible nucleotide substitution model with gamma-distributed rate variation among sites (GTR+Γ), combined with a strict molecular clock and a coalescent Bayesian skyline prior, provided the best fit to the data. Three independent MCMC chains were run, sampling parameters every 1,000 steps. Convergence and effective sampling were assessed using Tracer v1.8, ensuring that all parameters achieved effective sample size (ESS) values greater than 200. Log files from independent runs were combined using LogCombiner with a burn-in of 10%, and maximum clade credibility trees were generated using TreeAnnotator.

### Phylodynamic reconstruction

To investigate historical changes in bacterial population size, phylodynamic analyses were performed to estimate variation in effective population size (Ne) over time for major typhoidal *Salmonella* lineages circulating in Nepal. Time-scaled phylogenetic trees inferred from the Bayesian analyses were used to reconstruct lineage-specific effective population size trajectories using the skygrowth package (v0.3.1) in R. Separate reconstructions were performed for the H58 lineages, the lineage 3.3 group of *S.* Typhi, and the *S.* Paratyphi A population.

To assess whether the pandemic and post-vaccine declines were lineage-specific, we computed instantaneous per-lineage growth rates of the effective population size, r(t) = d log Ne/dt, from 100 randomly drawn time-calibrated trees from each BEAST posterior. Period-mean growth rates were summarized for three epidemiological windows (pre-pandemic, pandemic/pre-vaccine, and post-TCV) with 95% credible intervals derived from the posterior distribution, and the year of steepest decline was defined as the year with the minimum r(t). Cross-pathogen contrasts (Δr) were computed by subtracting the period-mean growth rate of *S.* Paratyphi A from each *S.* Typhi lineage in each period, with the posterior probability that the contrast was negative reported for each contrast. *S*. Paratyphi A, which is not targeted by current TCVs, served as a within-study negative control for vaccine-associated effects.

As an independent test for population bottleneck signatures, classical population-genetic statistics were computed for each lineage and each epidemiological period from the per-lineage SNP alignments. Nucleotide diversity (π) per site and Tajima’s D were calculated using scikit-allel v1.3.7. A drop in π coupled with a Tajima’s D shifting from strongly negative toward zero or positive values was interpreted as evidence of a recent population contraction; rising π with positive Tajima’s D was interpreted as diversification. These analyses were performed independently of the skygrowth reconstructions and used to triangulate the phylodynamic signal.

### Vi capsule and O-antigen variant analysis

To screen for evidence of vaccine-driven escape at the Vi capsule, all 350 Nepal *S*. Typhi isolates (200 pre-TCV, 150 post-TCV) were inspected at the 26 positions that were variant across the Vi biosynthesis (*tviA-E*, *vexA-E*), O-antigen biosynthesis (*rfbX/S/H*), and adjacent regulatory loci (*YncE*, *VirK1/2*). Variants were annotated at the codon level (synonymous, non-synonymous, or indel) against the CT18 reference and compared between the pre-TCV and post-TCV periods. Findings were independently verified by parsimony homoplasy and dN/dS analysis, and the screen was extended to 1,137 isolates comprising *S*. Typhi genomes from six TCV-introduction settings including India, Pakistan, Bangladesh, Malawi, Zimbabwe, and Nepal (S1 Table), screened at a pre-specified panel of 24 candidate Vi/O-antigen positions (spanning *rfbX*/*S*/*H*, *tviD*/*E*, and *vexC*/*E*) callable across all six settings to test reproducibility across endemic backgrounds.

## Results

### The population structure of *S*. Typhi and *S*. Paratyphi A isolates in Nepal

A total of 350 *S*. Typhi isolates were newly sequenced in this study, adding to previous genomic surveillance in Nepal. Among these, 168 (48%) were collected prior to the COVID-19 pandemic, 32 (9.1%) during the pandemic/pre-vaccine period, and 150 (42.9%) following the introduction of typhoid conjugate vaccines. Genotyping identified 16 distinct *S*. Typhi genotypes, indicating substantial genetic diversity within the population. The most common genotype was 4.3.1.2, accounting for 98 isolates (28%), followed by 3.3.2 with 71 isolates (20.3%) and 3.3.1 with 59 isolates (16.9%). The broader 4.3.1 lineage (H58) was represented by 42 isolates (12.0%), while additional sublineages included 4.3.1.2.1 (31 isolates, 8.8%) and 4.3.1.1 (6 isolates, 1.7%). Other genotypes were detected at lower frequencies. Among the 114 *S*. Paratyphi A isolates, 78 (68.4%) were collected prior to the pandemic and 36 (31.6%) during or after the pandemic period. Genotyping revealed seven distinct genotypes, with the population dominated by genotype 2.4.3, which accounted for 65 isolates (57.0%). The second most common genotype was 2.4.2 with 29 isolates (25.4%). Other genotypes were observed at substantially lower frequencies.

### Antimicrobial resistance profile

Most *S*. Typhi isolates (347/350, 99.1%) and all *S*. Paratyphi A isolates were genotypically predicted to be susceptible to the traditional first-line antibiotics co-trimoxazole, ampicillin, and chloramphenicol. Antimicrobial resistance genes associated with these antibiotics were detected in only three *S*. Typhi isolates, all of which exhibited a MDR profile. These isolates carried the resistance genes *blaTEM-1*, *catA1*, *dfrA7*, *sul1*, and *sul2*, and belonged to genotype 4.3.1.1. Reduced susceptibility to fluoroquinolones was common among both pathogens. Among *S*. Typhi, 247 isolates (70.6%) were non-susceptible to fluoroquinolones (FQNS), primarily due to mutations in the quinolone resistance-determining regions (QRDRs) of *gyrA*, *gyrB*, *parC*, and *parE*. Among the *S*. Typhi isolates, 31 (8.8%) carried the characteristic triple QRDR mutations (*gyrA* S83F, *gyrA* D87N, and *parC* S80I), which are associated with high-level fluoroquinolone resistance. All triple-mutant isolates belonged to the H58 lineage II (genotype 4.3.1.2.1). Among S. Paratyphi A, 113/114 isolates (99.1%) carried QRDR mutations.

When examined across the full Nepal genomic dataset, including both newly sequenced and previously published isolates, distinct temporal patterns in fluoroquinolone resistance were observed. In *S*. Typhi, isolates carrying single QRDR mutations predominated across most years, with triple-mutant isolates first detected in the sequenced sample after 2013 and remained a stable minority across the surveillance period (Fig 1A). Notably, the proportion of isolates without QRDR mutations increased in recent years, suggesting a partial re-emergence of fluoroquinolone-susceptible *S*. Typhi. This increase was statistically significant,with the proportion of isolates lacking QRDR mutations rising from 0.2% (2/1,138) in 2008-2018 to 28.3% (101/357) in 2019-2024 (Fisher exact test p < 0.001). In contrast, *S*. Paratyphi A showed persistently high levels of fluoroquinolone non-susceptibility across nearly the entire surveillance period, with only sporadic years in which susceptible isolates were observed (Fig 1B).

**Fig 1.**
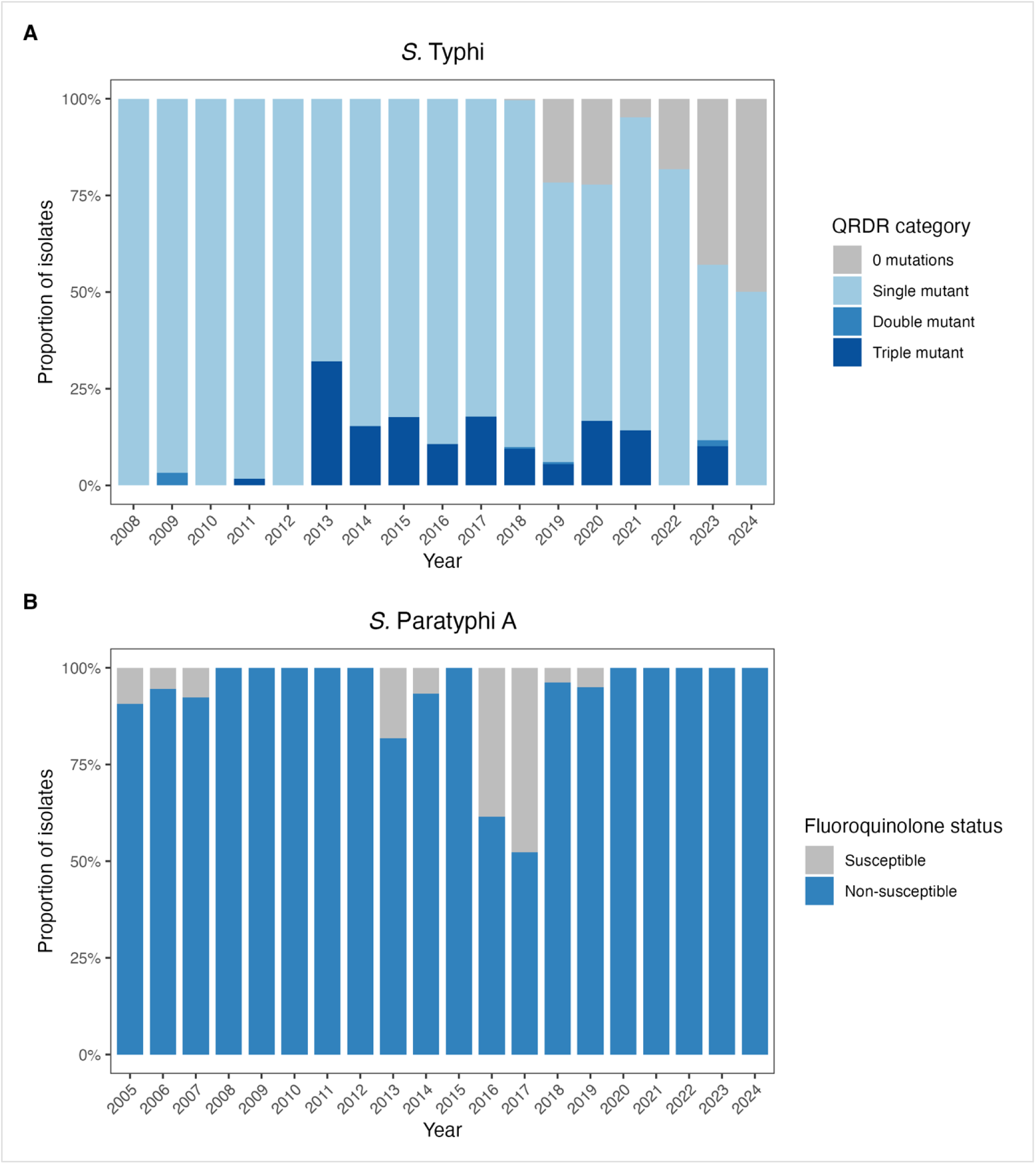
Temporal distribution of fluoroquinolone resistance-associated genotypes among typhoidal *Salmonella* isolates in Nepal. **(A)** Annual proportion of *S*. Typhi isolates classified according to the number of quinolone resistance-determining region (QRDR) mutations (0, 1, 2, or 3 mutations). **(B)** Annual proportion of *S*. Paratyphi A isolates classified as fluoroquinolone susceptible or non-susceptible based on the presence of QRDR mutations.

### Phylogenetic structure of typhoidal *Salmonella* in Nepal

To place recent Nepalese typhoidal *Salmonella* isolates in historical and phylogenetic context, we analyzed newly sequenced genomes alongside previously published genomes from Nepal. For *S*. Typhi, the contextual genomic dataset included 350 isolates generated in this study and 1,372 previously published genomes. For *S*. Paratyphi A, the corresponding dataset included 114 isolates generated in this study and 424 previously published genomes. This expanded Nepal dataset was used for phylogenetic reconstruction and for visualization of long-term temporal patterns in genotype and QRDR-associated fluoroquinolone resistance. Phylogenetic reconstruction demonstrated that both *S.* Typhi and *S*. Paratyphi A populations in Nepal were composed of multiple co-circulating lineages, but with clear dominance of a small number of successful genotypes. In *S.* Typhi, the phylogeny showed a broad distribution of isolates across several major clades, with H58-associated lineages forming a large proportion of the contemporary population and non-H58 lineages, including 3.3.1 and 3.3.2, remaining well represented throughout the study period (Fig 2). Within H58, time-calibrated Bayesian phylogenetic analysis of the full dataset further showed that the high-level fluoroquinolone-resistant clade (genotype 4.3.1.2.1, all carrying the gyrA-S83F + gyrA-D87N + parC-S80I triple QRDR mutation set) was strictly monophyletic, with a most recent common ancestor dated to 2008.2 (95% credible interval 2006.3-2009.9), consistent with a single ancestral acquisition of the third QRDR mutation followed by sixteen years of clonal expansion within Nepal rather than multiple independent emergences or recent importation (S1 Fig). In *S.* Paratyphi A, the phylogeny was more concentrated (Fig 3), with most isolates clustering within the 2.4 lineage, particularly genotype 2.4.3, consistent with a more restricted population structure compared with *S.* Typhi.

**Fig 2.**
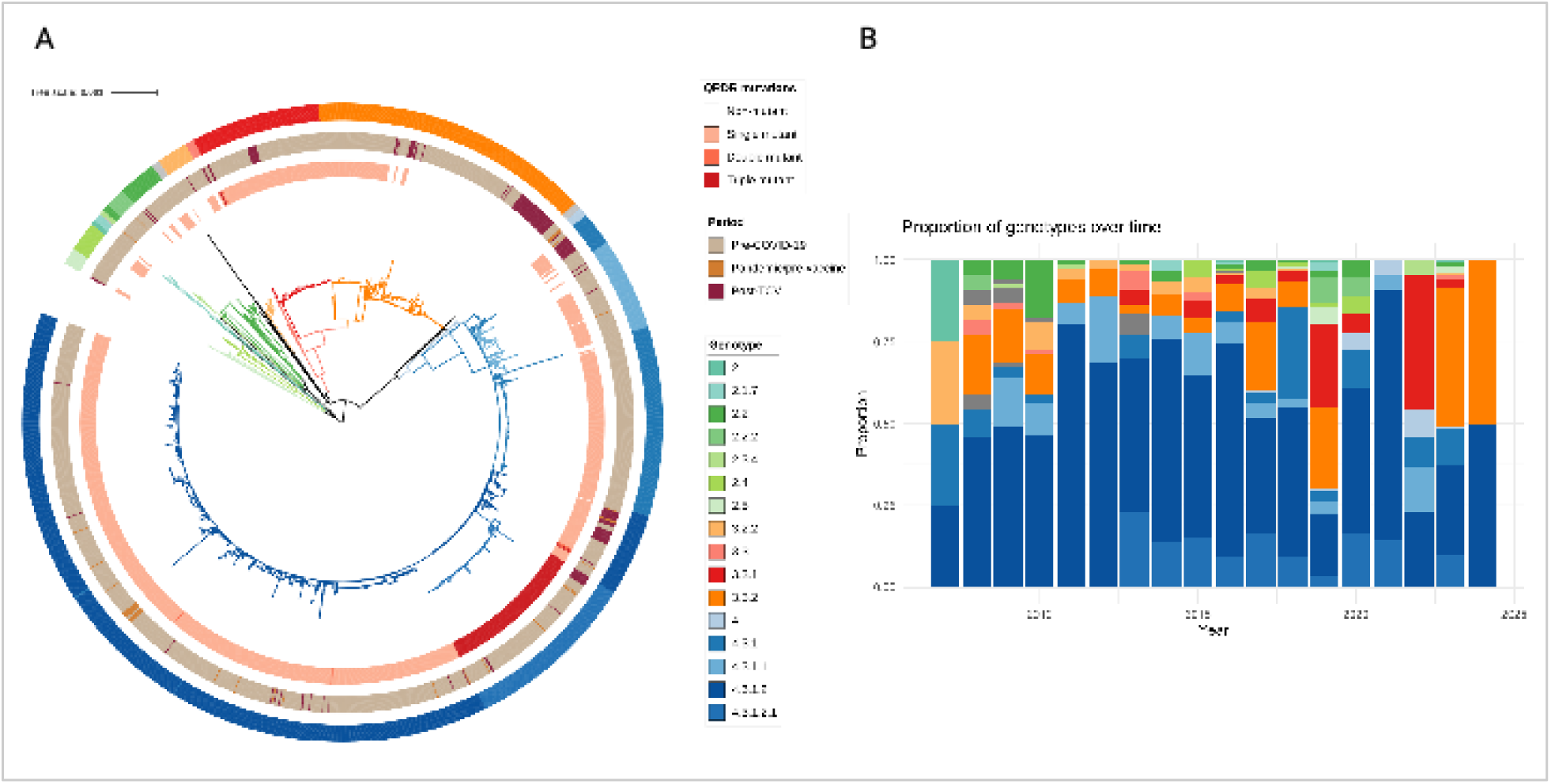
Phylogenetic structure and temporal distribution of *Salmonella* Typhi in Nepal. (**A**) Maximum likelihood phylogeny of 1,722 *S*. Typhi genomes from Nepal, including 350 isolates generated in this study and 1,372 previously published genomes. Branches and outer ring are colored by genotype according to the GenoTyphi scheme. Inner ring is colored according to the number of quinolone resistance-determining regions (QRDRs). The tree scale bar indicates nucleotide substitutions per site. (**B**) Temporal distribution of *S*. Typhi genotypes in Nepal. Stacked bar plots show the proportion of genotypes by year.

**Fig 3.**
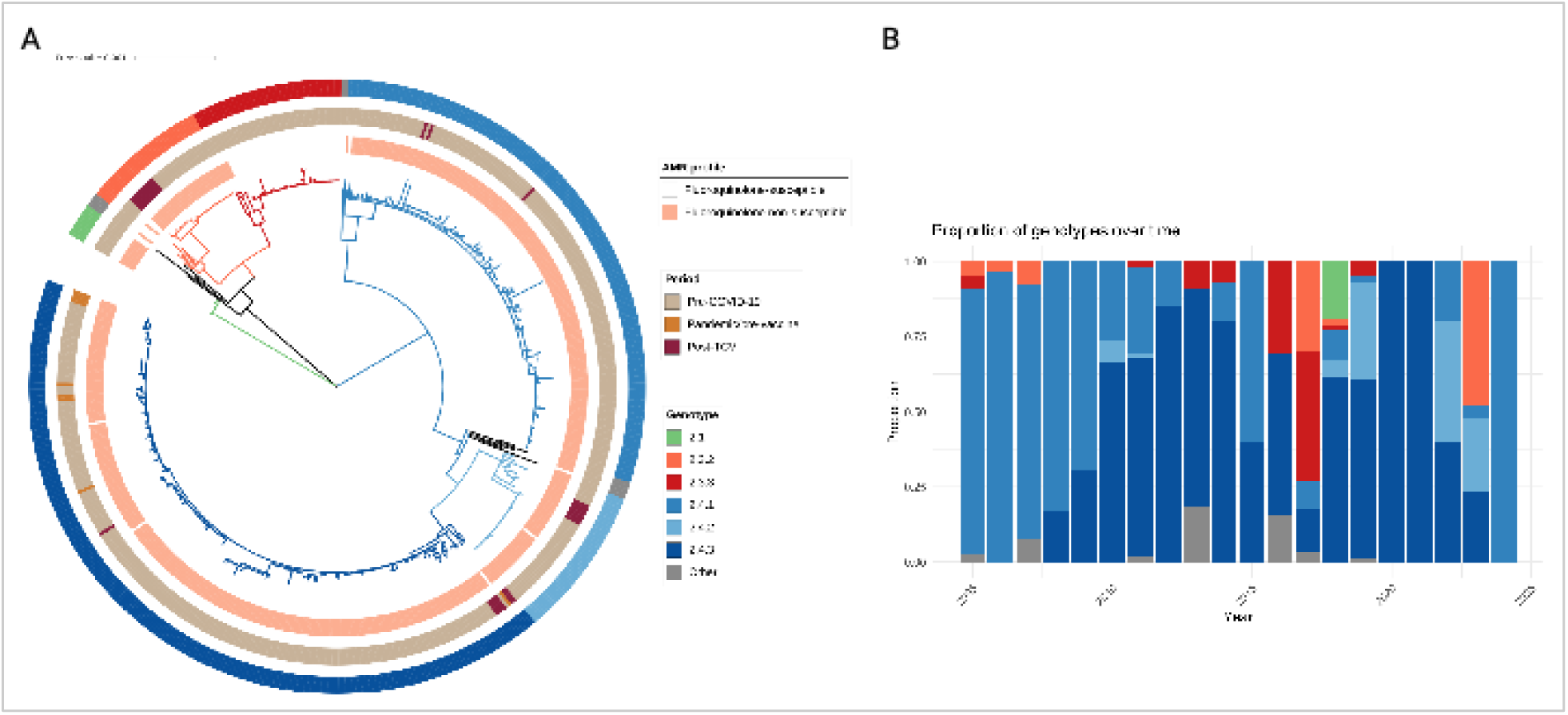
Phylogenetic structure and temporal distribution of Salmonella Paratyphi A in Nepal. (**A**) Maximum likelihood phylogeny of 538 *S*. Paratyphi A genomes from Nepal, including 114 isolates generated in this study and 424 previously published genomes. Branches and outer ring are colored by genotype according to the Paratype scheme. Inner ring is colored according to the antimicrobial resistance profile. The scale bar indicates nucleotide substitutions per site. (**B**) Temporal distribution of *S*. Paratyphi A genotypes in Nepal. Stacked bar plots show the proportion of genotypes by year.

### Temporal changes in effective population size of typhoidal *Salmonella* in Nepal

Phylodynamic reconstruction revealed distinct historical trajectories in the effective population size (Nₑ) of major lineages circulating in Nepal (S2 Fig). *S*. Typhi H58 (genotype 4.3.1) expanded from the early 2000s, peaked between 2014 and 2016, and contracted thereafter, with a markedly steeper decline following the pandemic onset in 2020 and a further drop after TCV introduction in 2022. Lineage 3.3 followed a different trajectory, declining gradually from the early 2000s and reaching low levels well before the pandemic or vaccine roll-out, with an additional sharp contraction emerging only after 2022. The combined *S*. Typhi profile integrated these dynamics, showing a clear two-step contraction aligned to the 2020 and 2022 inflection points. In contrast, *S*. Paratyphi A, was stable through the early 2010s, peaked around 2013– 2014, and declined gradually from 2016 onward, with no abrupt step-change coincident with either the pandemic or vaccine introduction.

### Lineage-specific population dynamics distinguish vaccine and pandemic effects

Per-lineage instantaneous growth rates derived from the posterior trees revealed strikingly different trajectories across the three major lineages (Fig 4). *S*. Typhi H58 contracted sharply during the pandemic (period-mean r =-0.63/year; steepest decline in 2021.5) and remained depressed without further change post-TCV. In contrast, lineage 3.3 was nearly flat during the pandemic but contracted sharply only after TCV introduction, reaching its steepest decline in 2023.0 (95% credible interval 2022.8-2023.2; period-mean r =-0.83/year). *S*. Paratyphi A, which is not targeted by the vaccine, declined modestly during the pandemic and showed no further change post-TCV. Using *S*. Paratyphi A as a within-study negative control, the post-TCV growth-rate difference was-0.43 for H58 (*p* = 0.92) and-0.70 for lineage 3.3 (*p* = 1.0), consistent with a lineage-3.3-specific, TCV-associated decline that is not seen in the non-vaccine-targeted comparator.

**Fig 4.**
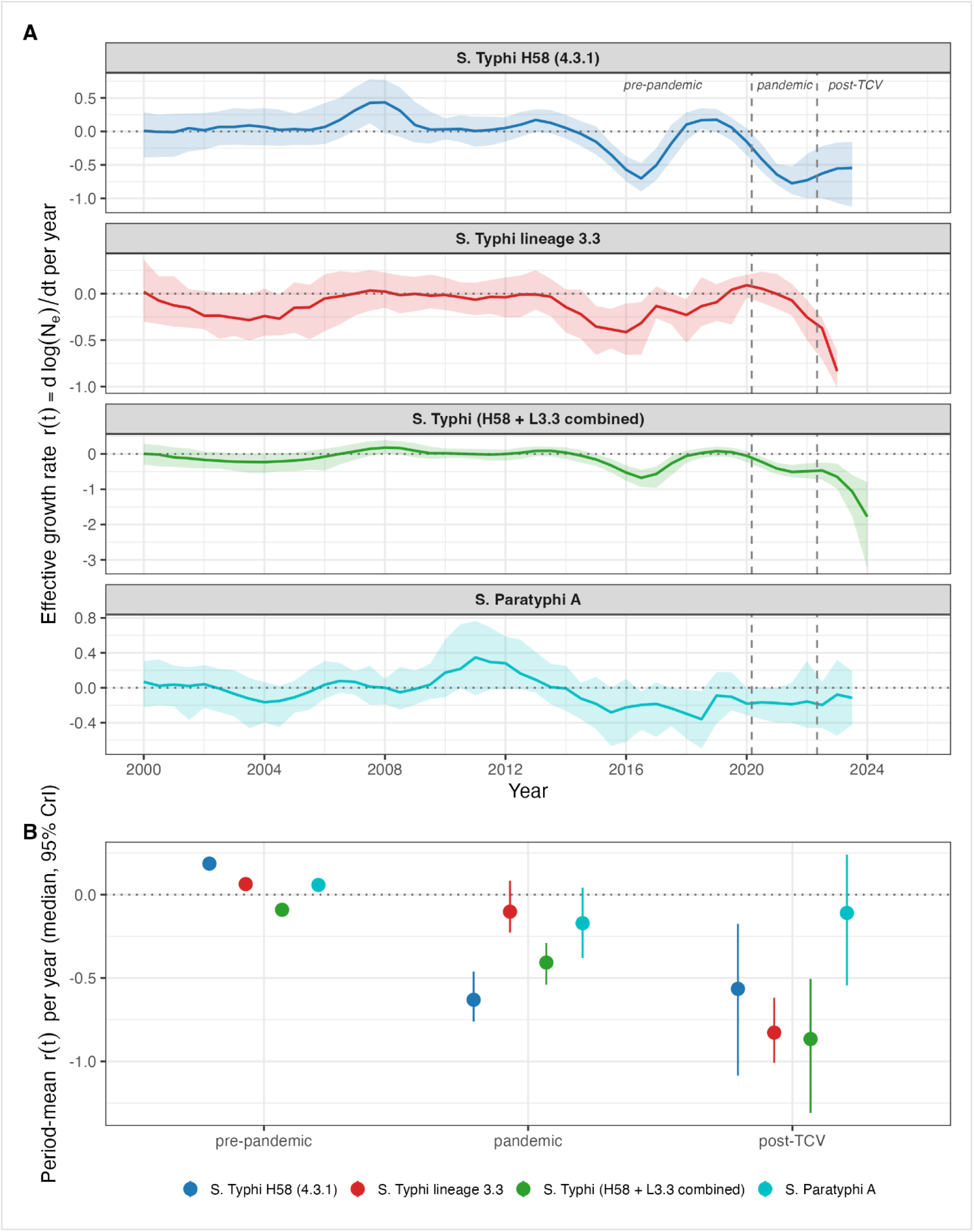
Differential lineage growth dynamics of S. Typhi and S. Paratyphi A in Nepal across the pre-pandemic, COVID-19 pandemic and post-TCV periods. Estimates are derived from a non-parametric skygrowth coalescent analysis applied to 100 BEAST posterior trees per lineage. **(A)** Instantaneous effective growth rate *r(t)* = d log *N*ₑ / d*t* per year, solid lines show the posterior median and shaded bands the 95% credible interval. Each panel shows one lineage: *S*. Typhi H58 - 4.3.1 (blue), *S*. Typhi lineage 3.3 (red), the combined H58 + lineage 3.3 *S*. Typhi population (green), and *S*. Paratyphi A (teal). The horizontal dotted line marks r(t) = 0 (constant Ne); vertical dashed lines mark the COVID-19 pandemic onset in Nepal (March 2020) and the national TCV rollout (April 2022) **(B)** Period-mean growth rate per lineage, summarized as the posterior median (point) and 95% credible interval (vertical bar) within each period.

Classical population-genetic statistics provided a third converging line of evidence (Table 1). *S*. Typhi lineage 3.3 showed the signature of a recent population contraction, with nucleotide diversity (π) dropping 36% in the post-TCV period. *S*. Typhi H58 showed no comparable loss of diversity post-TCV; instead, Tajima’s D drifted back toward zero, consistent with the population re-equilibrating after its pandemic-era contraction. In contrast, *S*. Paratyphi A moved in the opposite direction, with π rising by 34% and Tajima’s D shifting from-1.45 to +1.38, the signature of diversification rather than contraction.

**Table 1.** Population-genetic diversity statistics by lineage and epidemiological period. Nucleotide diversity (π) per site and Tajima’s *D* were computed from per-lineage SNP alignments. Pandemic = April 2020-March 2022; Post-TCV =April 2022-March 2024. Percentages in parentheses indicate change in π relative to the pre-pandemic baseline.

| Lineage | Period | n | $\pi$ / site | Tajima's $D$ |
| --- | --- | --- | --- | --- |
| ***S. Typhi* H58 (4.3.1)** | Pre-pandemic | 305 | 0.0195 | -2.53 |
|  | Pandemic | 23 | 0.0183 | -1.93 |
|  | Post-TCV | 31 | 0.0232 (+19%) | -0.29 |
| ***S. Typhi* lineage 3.3** | Pre-pandemic | 308 | 0.0708 | -1.57 |
|  | Post-TCV | 68 | 0.0453 (-36%) | -1.35 |
| ***S. Paratyphi* A** | Pre-pandemic | 503 | 0.00214 | -1.45 |
|  | Pandemic | 9 | 0.00016 | -1.31 |
|  | Post-TCV | 27 | 0.00286 (+34%) | +1.38 |

### Signatures of TCV impact on the age and genotype distribution

The age distribution of *S.* Typhi cases shifted older following TCV introduction. Median age increased from 19 years pre-TCV to 22 years post-TCV (Mann-Whitney *p* = 9.3 × 10⁻⁵), and the proportion of cases in the vaccine-eligible age group (15 months to 15 years) decreased from 31% to 14% (Fisher exact *p* = 2.1 × 10⁻⁴), providing direct evidence of vaccine impact on the targeted age group (S3A Fig). Among unvaccinated adults (>15 years), the overall genotype composition differed significantly between periods (three-class χ² *p* = 0.005). The share of lineage 3.3 increased from 33% to 47% (Fisher exact *p* = 0.024) and minor genotypes decreased from 17% to 5% (Fisher exact *p* = 0.004), while the H58 share was essentially unchanged (S3B Fig). District-stratified analysis (S2 Table) showed that the post-TCV shift in *S*. Typhi composition was concentrated in Kathmandu, where lineage 3.3 rose from 41.5% (pre-TCV) to 53.0% (post-TCV) of cases and minor genotypes reduced from 17.0% to 5.1%, while H58 (4.3.1) remained near 42%. *S*. Paratyphi A showed a marked decline of genotype 2.4.3 in Kathmandu (63.3% pre-TCV vs. 10.0% post-TCV) with a rise of minor genotypes, although the small post-TCV *S*. Paratyphi A sample across districts warrants cautious interpretation.

### No evidence of vaccine-driven escape at the Vi capsule or O-antigen

Variant scanning of the Vi capsule and O-antigen biosynthesis loci provided no evidence of vaccine-driven escape. No non-synonymous variant was exclusive to post-TCV isolates; the only premature stop codon identified (*tviD* C798*) was carried by three pre-TCV genotype 4.3.1.2.1 isolates, and the two synonymous variants showed period-associated frequency changes in opposite directions, consistent with sub-lineage replacement rather than vaccine-driven selection. In the 1,137-isolate cross-country cohort spanning five additional TCV-introduction settings, only a single synonymous variant reached pooled significance (*rfbH* K263K), likely reflecting subclade population structure rather than functional adaptation. In parallel, the Vi-loss screening analysis identified no confirmed Vi-negative *S*. Typhi isolates.

## Discussion

In this study, we combined prospective genomic surveillance with historical datasets to characterize long-term changes in the population structure and antimicrobial resistance profiles of *S*. Typhi and *S*. Paratyphi A in Nepal, and to assess how the size and composition of these bacterial populations evolved across three key epidemiological periods: before the COVID-19 pandemic, during the pandemic, and following the introduction of typhoid conjugate vaccines (TCVs). Our findings reveal a marked recent decline in the effective population size of both pathogens, lineage-specific compositional shifts, and persistently high levels of fluoroquinolone resistance. Together, these results provide new insight into how major public health interventions and large-scale social disruptions may influence patterns of typhoidal *Salmonella* circulation in endemic settings.

Phylodynamic analyses revealed a consistent pattern of population expansion followed by decline across major lineages, but the timing of these declines was asymmetric in a way that allows the contributions of the pandemic and TCV introduction to be partially separated. *S*. Typhi H58 contracted sharply during the pandemic and remained depressed thereafter, whereas lineage 3.3 was nearly flat during the pandemic and reached its steepest decline only post-TCV. *S*. Paratyphi A, which is not targeted by the vaccine, showed only a modest pandemic-era decline and no further change post-TCV, providing a within-study negative control for vaccine-associated effects. This pattern was independently corroborated by classical population-genetic statistics: nucleotide diversity in lineage 3.3 dropped 36% post-TCV, H58 equilibrated after its pandemic-era contraction, and *S*. Paratyphi A diversified rather than contracting. Together, these three independent genomic lines of evidence (per-lineage growth-rate trajectories, cross-pathogen contrasts, and population-genetic diversity statistics) suggest that the pandemic-era decline was most pronounced in the most actively transmitting lineage (H58), whereas the post-TCV decline was concentrated in lineage 3.3. Although observational genomic data cannot establish causality directly, the contrasting trajectories between vaccine-targeted *S*. Typhi lineages and non-targeted *S*. Paratyphi A are consistent with TCV contributing to the observed post-2022 lineage 3.3 contraction.

These trends are consistent with emerging epidemiological data from Nepal and other endemic settings documenting substantial reductions in typhoid incidence during the COVID-19 pandemic [9,10,35,36]. The post-vaccine period, however, was more heterogeneous: clinical surveillance in Nepal documented continued declines among vaccine-eligible children but a concurrent rise among adults, who comprise the majority of cases, such that overall typhoid incidence rose following 2022 [10]. Our findings extend these observations by demonstrating parallel declines in the effective population size of both *S*. Typhi and *S*. Paratyphi A, providing genomic evidence supporting a true reduction in pathogen circulation rather than changes in surveillance or healthcare utilization alone. The post-TCV genotype shift observed among unvaccinated adults represents an indirect, herd-mediated signal of vaccine impact, in which the genotype pool to which the unvaccinated adult population is exposed is reshaped by the suppression of transmission in the vaccinated paediatric age group [6,7]. Our findings are consistent with the published TCV-effectiveness studies. Randomized trials in Malawi [6] and Bangladesh [7], a programmatic evaluation in Navi Mumbai [8], and a recent real-world meta-analysis [9] have all documented substantial reductions in paediatric typhoid following TCV introduction, while parallel genomic surveillance from Zimbabwe has shown contraction of the dominant H58 lineages after vaccine rollout [35]. Our data extend this landscape by demonstrating that, even when TCV was deployed immediately after a major pandemic-era contraction, an additional post-TCV signal remained detectable at the genomic level, and was lineage-specific, supporting the generalizability of TCV impact across diverse endemic backgrounds.

The decline in *S*. Paratyphi A, which is not targeted by current vaccines [11], suggests that factors beyond vaccination likely contributed to reduced transmission. These include pandemic-associated behavioral and environmental changes, such as reduced population mobility, altered healthcare-seeking behavior, and improvements in hygiene practices during the COVID-19 pandemic [3]. A genotype-level analysis restricted to Kathmandu, where most isolates were sampled, further suggested a marked shift in the *S*. Paratyphi A genotype mix in the post-TCV period, with the historically dominant genotype 2.4.3 contracting and minor genotypes accounting for the majority of recent isolates. Whether this reflects a true compositional turnover or sampling variation in a declining population will require longer follow-up. However, we did not observe clear evidence of serovar replacement, as the overall *S*. Paratyphi A population itself also contracted, indicating that vaccination has not produced an immediate expansion of the non-vaccine serovar. Sustained circulation of fluoroquinolone-non-susceptible *S*. Paratyphi A raises concern that successful typhoid control could progressively unmask paratyphoid fever as a proportionally larger enteric fever burden. Continued surveillance will be important to monitor longer-term changes in serovar distribution and to inform the case for a bivalent conjugate vaccine targeting both serovars [11].

A central concern when introducing a polysaccharide-conjugate vaccine into an endemic setting is the possibility of vaccine-driven escape at the antigenic target [6,7]. We found no non-synonymous variant exclusive to post-TCV isolates and no confirmed Vi-loss isolates in any setting. The absence of detectable vaccine escape across endemic backgrounds, together with the strict monophyly and 2008 origin of the high-level fluoroquinolone-resistant clade, provides reassurance that the post-TCV residual transmission reflects local persistence of the existing population under vaccine pressure rather than the emergence of an antigenically distinct escape lineage.

The results of this study should be interpreted within the context of some limitations. First, the number of isolates collected during the pandemic period was relatively small, which may limit the precision of temporal inferences. Second, our dataset is largely derived from population-based surveillance in specific geographic regions, which may not fully capture transmission dynamics across Nepal. Third, district-stratified analyses are anchored on Kathmandu, which dominates the cohort; signals in peri-urban and rural districts rest on small numbers of isolates, particularly in the post-TCV period for *S*. Paratyphi A. Fourth, phylodynamic estimates of effective population size are influenced by sampling density and may not directly reflect true incidence. Finally, although our cross-pathogen contrasts using *S*. Paratyphi A as a within-study negative control provide a partial causal anchor for the asymmetric vaccine-versus-pandemic interpretation, formal causal inference will require longer post-TCV follow-up and ideally individual-level vaccine histories.

In conclusion, our integrated genomic and phylodynamic analysis demonstrates that both *S*. Typhi and *S*. Paratyphi A populations in Nepal have undergone substantial recent declines in effective population size, alongside lineage-specific compositional shifts, and widespread fluoroquinolone resistance. These findings highlight the potential impact of vaccination and large-scale social disruptions on enteric fever epidemiology and underscore the importance of sustained genomic surveillance. Expanding vaccine strategies to include *S*. Paratyphi A and strengthening antimicrobial stewardship can contribute importantly to long-term control of enteric fever.

## Data sharing

Raw sequence data generated in this study was submitted to the European Nucleotide Archive under the study accession number PRJEB121301. Sequence data from previous studies were available in European Nucleotide Archive. Details and individual accession numbers of sequence data included in our analysis have been included in S3 Table and S4 Table.

## Ethics statement

This study received ethical approval from the Nepal Health Research Council and the Kathmandu University School of Medical Sciences. Written informed consent was obtained from participants or their guardians prior to study enrollment. Confidentiality of participant data was ensured by de-identifying all records and aggregating blood culture results prior to analysis.

## Data Availability

Raw sequence data generated in this study was submitted to the European Nucleotide Archive under the study accession number PRJEB121301. Sequence data from previous studies were available in European Nucleotide Archive. Details and individual accession numbers of sequence data included in our analysis have been included in Table S1 and Table S2.

https://www.ebi.ac.uk/ena/browser/view/PRJEB121301

## Acknowledgments

This study was funded by the Gates Foundation (grant INV-008335 to D.O.G.; https://www.gatesfoundation.org). The funders had no role in study design, data collection and analysis, decision to publish, or preparation of the manuscript.

## Supporting information

S1 Fig. Posterior distribution of the most recent common ancestor (MRCA) date for the Nepal high-level fluoroquinolone-resistant (triple-QRDR) S. Typhi 4.3.1.2.1 clade. MRCA dates were estimated across 100 trees sampled from the BEAST posterior. The upper panel shows the MRCA of all (n=156) 4.3.1.2.1 isolates in the H58 phylogeny; the lower panel restricts to the 4.3.1.2.1 isolates newly sequenced in our Nepal study (n=31), all carrying the triple-QRDR genotype (gyrA-S83F + gyrA-D87N + parC-S80I). Vertical dashed lines mark the onset of the COVID-19 pandemic in Nepal (March 2020) and the national TCV rollout (April 2022).

S2 Fig. Effective population size Ne(t) trajectories per lineage from a skygrowth analysis of dated BEAST posterior trees. Each panel shows the posterior median Ne(t) (solid line) with the 95% credible interval (shaded band) on a log10 scale. Lineages: S. Typhi H58 / 4.3.1 (blue), S. Typhi lineage 3.3 (red), the combined H58 + lineage 3.3 S. Typhi population (green), and S. Paratyphi A (teal). Vertical dashed lines mark the start of the COVID-19 pandemic in Nepal (March 2020) and the national TCV rollout (April 2022).

S3 Fig. Direct and indirect signatures of typhoid conjugate vaccine (TCV) impact on the S. Typhi case population in Nepal. (A) Age distribution of sequenced S. Typhi cases before and after TCV introduction; the shaded blue band and dotted vertical lines mark the vaccine-eligible age window (15 months – 15 years). (B) S. Typhi lineage composition by host age class, before and after TCV: H58 / 4.3.1 (blue), lineage 3.3 (red), and other genotypes (orange).

S1 Table. Multi-country Salmonella Typhi genomic cohort used to test the generalizability of the Nepal post-TCV findings.

S2 Table. Within-Nepal district patterns of Salmonella Typhi and Salmonella Paratyphi A genotypes, before and after programmatic TCV introduction (April 2022).

S3 Table. Genomic metadata and European Nucleotide Archive accession numbers for the 350 newly sequenced S. Typhi isolates from this study.

S4 Table. Genomic metadata and European Nucleotide Archive accession numbers for the 114 newly sequenced S. Paratyphi A isolates from this study.

